# Efficacy and safety of hydroxychloroquine for the treatment of osteoarthritis: protocol for a systematic review of randomized controlled trials

**DOI:** 10.1101/2020.07.20.20157669

**Authors:** Ambrish Singh, Anirudh Kotlo, Thusharika Dissanayaka, Zhiqiang Wang, Benny Antony

## Abstract

Hydroxychloroquine (HCQ) is a conventional disease-modifying antirheumatic drug (DMARD), which is considered as relatively safe, and offers a modest efficacy profile for the treatment of inflammatory rheumatic diseases such as rheumatoid arthritis and systemic lupus erythematosus. In view of the anecdotal evidence on its immunomodulatory and anti-inflammatory properties, HCQ has been used as an off-label option in patients with osteoarthritis (OA), mainly for the treatment of inflammatory OA. Recently, many investigators have evaluated the safety and efficacy of HCQ for the treatment of OA in various randomized control trials (RCTs). While most RCTs have evaluated the HCQ in inflammatory OA (erosive hand OA), there are studies constituting knee OA patients as well. Currently, there are no systematic reviews that have summarized the evidence on the efficacy and safety of HCQ in OA population. Hence, this study aims to systematically review the evidence from RCTs assessing the efficacy and safety of HCQ for the treatment of OA. Biomedical databases such as PubMed, Embase, and Google Scholar will be searched to identify the RCTs of HCQ in patients with OA (hand, knee, hip, or any other OA). Cochrane risk of bias tool will be used to assess the quality of included studies. Review Manager 5 (Rev Man) and STATA Version 16 will be used to conduct the statistical analysis.

## 1. Background and rationale

Osteoarthritis (OA) is the most common type of arthritis that mainly affects the joints of knees, hips, hands, and feet. OA is characterized by inflammation and progressive deterioration of articular cartilage, synovium, ligaments, and the subchondral bone.(1) The epidemiological burden of OA increases with age and is more prevalent in women compared with men.(2) Currently, there is no disease-modifying treatment available for OA, and the current management options such as nonsteroidal anti-inflammatory drugs (NSAIDs) mostly offer only moderate symptomatic relief and have a poor safety profile. Hydroxychloroquine (HCQ), a conventional disease-modifying antirheumatic drug (DMARDs) characterized by its immunomodulatory and anti-inflammatory properties has been an effective treatment option for autoimmune diseases for many years. Furthermore, various studies have elucidated the benefits of HCQ emphasizing its favorable safety profile and outcomes in rheumatoid arthritis (RA) patients.(3) It has also been occasionally used as an off-label option, in erosive hand OA population.(4) Considering inflammation as one of the key features of OA and promising efficacy of conventional DMARDs in inflammatory arthritis such RA, recently many randomized controlled trials (RCTs) have evaluated HCQ in patients with OA.(5) However, the effectiveness of HCQ in patients with OA is now increasingly being questioned mostly due to the negative findings from RCTs in different OA population (such as hand and knee OA). Hence we plan to conduct a systematic review of evidence from RCT assessing the efficacy and safety of HCQ in patients with any type of OA (hand, knee, hip).

## 2. Objectives

The objective of this protocol is to outline the plan for the conduct of a systematic review and possible meta-analysis of RCTs assessing the safety and efficacy of HCQ for the treatment of OA.

## 3. Methodology

### 3.1 Eligibility criteria

We plan to select eligible studies based on specific criteria for Participants, Interventions, Comparators, Outcomes, and Study design (PICOS) We will conduct and report this systematic review in accordance with Preferred Reporting Items for Systematic Reviews and Meta-Analyses (PRISMA) statement.(6) We will include the studies published in the English language.

### 3.2 Characteristics of participants (P)

Adult participants aged at least 40 years old, of any sex and with a confirmed diagnosis of any type of OA (knee, hand, hip) according to the American College of Rheumatology or similar will be included as the population of interest.(7)

### 3.3 Characteristics of interventions (I)

The studies assessing the safety and efficacy of oral HCQ at any dose, commonly known with the brand name as Plaquenil (or any other brand), will be included.

### 3.4 Characteristics of comparators (C)

Studies comparing HCQ with placebo or any other active pharmacological comparator (such as, but not limited to, NSAIDs) for the treatment of OA will be included. All other non-pharmacological interventions such as exercise, acupuncture, physiotherapy, occupational and psychosocial therapy will be excluded

### 3.5 Characteristics of outcomes (O)

The studies reporting data for at least one of the following outcomes: OA related pain intensity [assessed using visual analog scale (VAS) score, Western Ontario and McMaster Universities Osteoarthritis Index (WOMAC) Score AUSCAN, NRS, or any other scale], quality of life (QoL) [assessed using EQ-5D, SF-6D or other scales], and safety will be included in the analysis.

### 3.6 Characteristics of study design (S)

We will include randomized, quasi-randomized, controlled, blinded, or open-label study only. Observational and non-randomized studies will be excluded.

## 4. Outcomes of interest

### 4.1 Efficacy outcomes

Our efficacy outcomes of interest will be

1. the change in OA associated pain intensity [assessed using VAS, WOMAC, AUSCAN, NRS, KOOS, and other scales]
2. the change in OA associated physical dysfunction [assessed using VAS, WOMAC, AUSCAN, NRS, KOOS, and other scales]
3. QoL [assessed with EQ-5D, SF-6D, HAQ or other scales], radiographic structural damage, and biomarker change.

### 4.2 Safety outcomes

Safety outcomes will include adverse effects/events reported with HCQ.

## 5. Information sources and search procedure

The search procedure will be employed in accordance with the following criteria:

### 5.1 Electronic source and search strategy

We will search PubMed, Embase, and Cochrane Central Register of controlled trials from inception to June 2020 using the below search strategy.

**Table 1:**
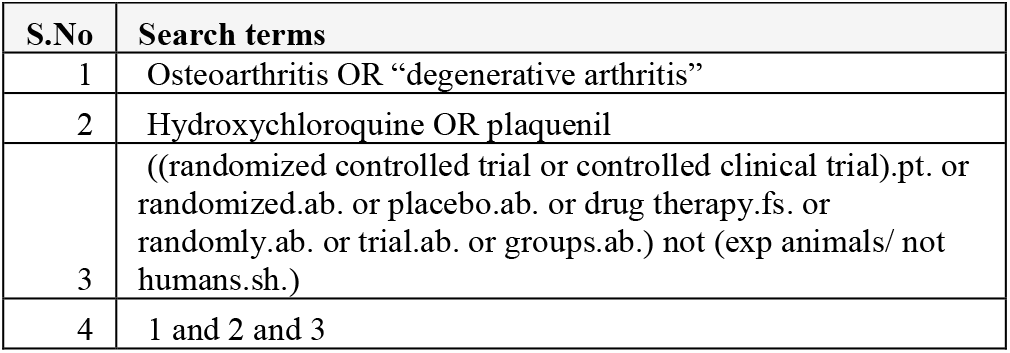
Representative search strategy.

| S.No | Search terms |
| --- | --- |
| 1 | Osteoarthritis OR "degenerative arthritis" |
| 2 | Hydroxychloroquine OR plaquenil |
| 3 | ((randomized controlled trial or controlled clinical trial).pt. or randomized.ab. or placebo.ab. or drug therapy.fs. or randomly.ab. or trial.ab. or groups.ab.) not (exp animals/ not humans.sh.) |
| 4 | 1 and 2 and 3 |

Bibliography of the included studies, relevant systematic reviews, meta-analysis, and Google Scholar will be searched to identify any relevant eligible trials.

### 5.2 Hand-searching

We will search the abstracts from last two years conference proceedings of major international association involved in OA research such as European League Against Rheumatism (EULAR), Osteoarthritis Research Society International (OARSI), American Academy of Orthopedic Surgeons (AAOS), and American College of Rheumatology (ACR).

### 5.3 Study selection

Two reviewers will independently screen the retrieved citations first based on title and abstract, then for the included citation based on full-text. The discrepancy in inclusion/exclusion of the studies will be resolved by mutual discussion or arbitration by the third reviewer.

### 5.4 Assessment of risk of bias

Cochrane risk of bias (RoB) tool will be used for the evaluation of the methodological quality of the included studies.(8) Two Researchers will independently evaluate each study using the RoB tool. The Cochrane RoB tool assess study characteristics reported for the following items: sequence generation, allocation concealment, blinding of participants, study personnel and outcome assessors, incomplete outcome data, selective outcome reporting, and other potential sources of bias. Any conflicts would be resolved by discussion.

## 5. Data extraction

Data would be extracted independently from included studies by two reviewers for the following information: study design, characteristics of the population (age, sex, BMI, and type of OA), sample size, intervention details and dosage, duration of follow-up, type of placebo or other control, outcome measurements, mean change values of the relevant outcome, the number of adverse events reported, and medication change. Data will be extracted for baseline and follow up values or mean change from baseline for each outcome of interest. Wherever applicable intention- to- treat data will be used. Corresponding authors may be contacted for key information when data deemed to be ambiguous or missing. Any discrepancy in data extraction will be resolved by mutual discussion or arbitration by the third reviewer.

## 6. Statistical analysis

We expect heterogeneity among included studies, due to different types of OA population and outcomes assessed, to be appropriate for meta-analysis. However, if data permits we will perform the meta-analysis of the included studies using the random effect model. Statistical heterogeneity will be assessed as per Q statistics (P<0.05 was considered heterogeneous) and I^2^ statistic (I^2^ >50% will be considered heterogeneous).(9) All statistical analyses will be performed using STATA version 16 (STATA Corp., Texas, USA) and Review Manager 5 (RevMan 5.3) (Copenhagen: The Nordic Cochrane Centre, The Cochrane Collaboration, 2014).

## Data Availability

Not applicable

